# Redefining the Bladder Cancer Phenotype using Patterns of Familial Risk

**DOI:** 10.1101/19003681

**Authors:** Heidi A. Hanson, Claire L. Leiser, Christopher Martin, Sumati Gupta, Ken R. Smith, Christopher Dechet, William Lowrance, Brock O’Neil, Nicola J. Camp

## Abstract

Relatives of bladder cancer (BCa) patients have been shown to be at increased risk for kidney, lung, thyroid, and cervical cancer after correcting for smoking related behaviors that may concentrate in some families. We demonstrate a new method to simultaneously assess risks for multiple cancers to identify distinct multi-cancer configurations (multiple different cancer types that cluster in relatives) surrounding BCa patients. We identified 6,416 individuals with urothelial carcinoma and familial information using the Utah Cancer Registry and Utah Population Database (UPDB). First-degree relatives, second-degree relatives, and first cousins were used to construct a familial enrichment matrix for cancer-types previously shown to be individually associated with BCa. K-medioids clustering were used to identify Familial Multi-Cancer Configurations (FMC). A case-control design and Cox regression with a 1:5 ratio of BCa cases to cancer-free controls was used to quantify the risk in specific relative-types and spouses in each FMC. Clustering analysis revealed 12 distinct FMCs, each exhibiting a different pattern of cancer co-aggregation. Of the 12 FMCs, four exhibited strong familial risk of bladder cancer along with specific patterns of increased risk of cancers in other sites (BCa FMCs), and were the focus of further investigation. Cancers at increased risk in these four BCa FMCs most commonly included melanoma, prostate and breast cancer and less commonly included leukemia, lung, pancreas and kidney cancer. A network-based approach can be used with familial data to discover new phenotype clusters for BCa, providing new directions for discovering patterns of cancer clustering.

## Introduction

Relatives of bladder cancer (BCa) patients are at increased risk for kidney, lung, thyroid, and cervical cancer after correcting for smoking related behaviors (1-4). Family history is an important risk factor for many cancers, which may extend across cancer types (5, 6). Simultaneously assessing risks to multiple cancers and identifying multi-cancer configurations (multiple cancer types that cluster in relatives) surrounding BCa will help elucidate complexities of shared cancer risk across organ sites, such as genetics or environmental exposures. Understanding multi-cancer configurations involving BCa is important for identifying shared risk factors and for counseling families of BCa patients about risk of other cancers.

Breast cancer (BrCa) is an exemplar for multi-cancer configurations. Approximately 30% of hereditary BrCa is explained by intermediate and high-risk inherited variants, like BRCA1 and BRCA2. Key factors contributing to the discovery of BRCA1 were dense familial clustering, early age of onset, and coaggregation with ovarian cancer (7-9). Unique multi-cancer configurations for carriers of BRCA1 mutations (breast, ovarian, Fanconi anemia, prostate, pancreatic, fallopian tube and peritoneal cancers) and BRCA2 mutations (breast, male breast, prostate, pancreatic cancers and Fanconi anemia) are now accepted (Figure 1). These multi-cancer configurations were identified after discovery of BRCA1/2 mutations in BrCa. However, data driven methods make it possible to uncover multi-cancer configurations before gene discovery. These configurations could immediately permit better definitions of cancer phenotypes (subtypes or multi-cancer) for gene discovery and environmental risk factor studies.

**Figure 1.**
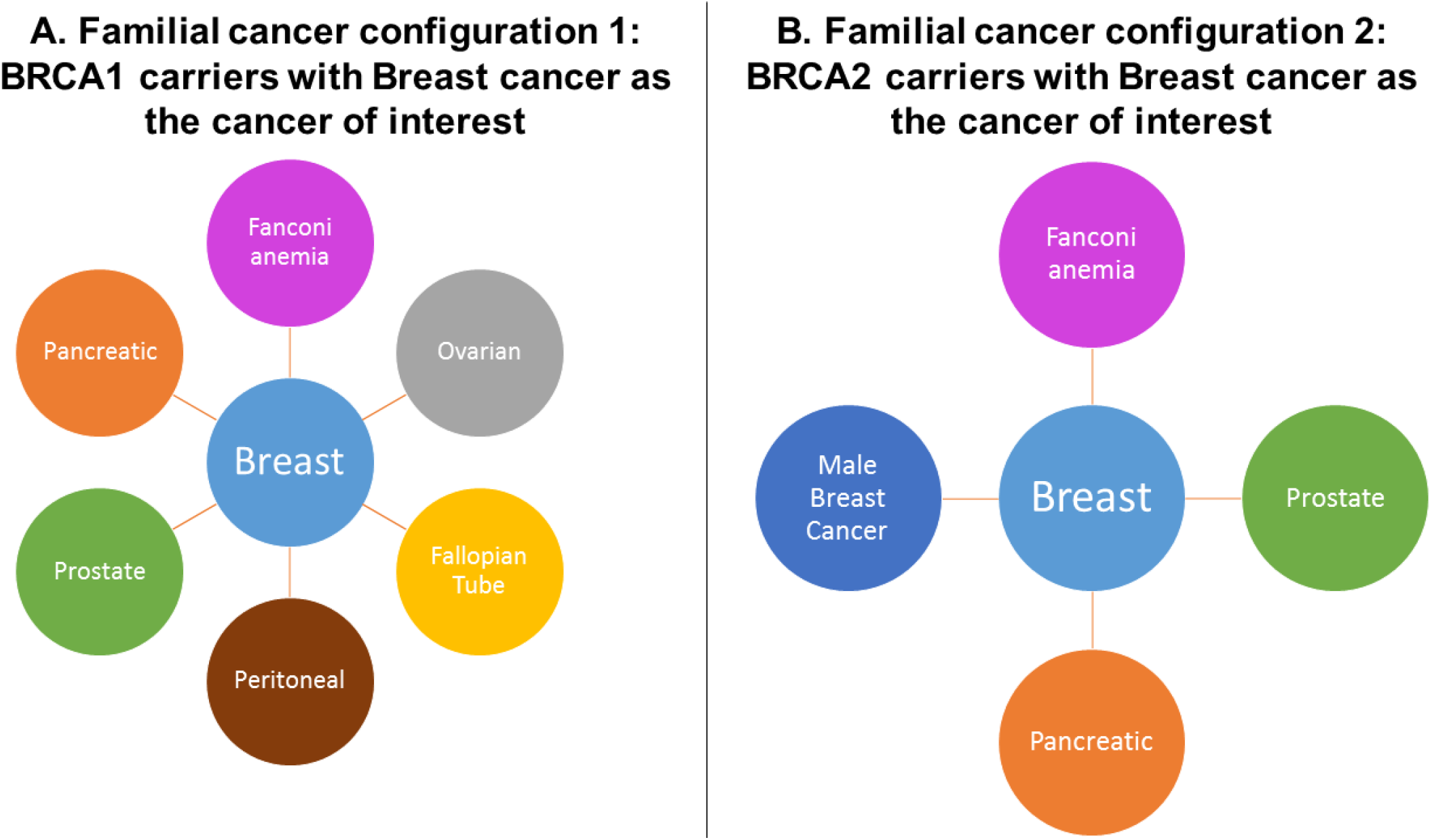
The familial multi-cancer configurations known to occur with subtypes of breast cancer (pivot). Panels A and B show familial multiphenograms of the known relationship with BRCA1 and BRCA2 genes, respectively.

Classical methods for assessing familial coaggregation are to determine the relative risk of cancer in first-degree (FDR), second-degree (SDR), and third-degree (TDR) relatives of individuals with the cancer of interest (pivot-individual). Cancer in the pivot-individual and cancer assessed in the relatives can be equivalent or different(10, 11). However, these methods use an iterative pairwise approach (pivot cancer and relative cancer) and are unable to simultaneously exploit data for multiple cancer types so power to identify novel patterns of multi-cancer clustering is limited. Other domains have developed innovative computational techniques to discover patterns across expansive amounts of data in large databases. Application of such “big-data” techniques to linked genealogical and cancer databases can potentially identify new multi-cancer configurations and improve understanding of familial cancer risk, tumor spectrum and phenotypic heterogeneity.

This study takes advantage of a unique population-level data resource, the Utah Population Database (UPDB), containing vast genealogy and statewide cancer data. This study focuses on the discovery of novel familial multi-cancer configurations for BCa.

## Methods

### Study design and data

We utilized the genealogical, demographic, and health data from the UPDB. The UPDB supports hundreds of biodemographic, epidemiologic, and genetic studies primarily due to its comprehensive population coverage, pedigree complexity, and linkages across data sources (12, 13).

Cancer-specific data for individuals with urothelial BCa and their relatives was obtained from the Utah Cancer Registry (UCR), an original member of the Surveillance Epidemiology and End Results (SEER) program. We identified 6,752 individuals born 1900-1990 with urothelial BCa and family history information (defined as at least ten known relatives) as pivots. Families were only represented once in the analysis. When multiple siblings (n ≥ 2) had BCa (nsibsets = 104), one sibling was randomly selected as a pivot. Our final sample included 6,416 three-generation families.

Sixteen cancer-types previously shown to be associated with BCa (thyroid, leukemia, pancreas, brain/CNS, lung, stomach, liver, larynx, melanoma, prostate, breast, ovarian, kidney, myeloma, small intestine, and ocular) (1, 14), plus BCa (17 cancers total) were considered simultaneously.

### Statistical Analysis

We constructed a 6,416 × 17 matrix and used k-medoids clustering(15) to measure similarities between the cancer enrichment profiles of families. Each resulting cluster was a BCa familial multi-cancer configuration (FMC).

### Familial Enrichment

The following histologies were not included (8000, 8001, 8010, 8041, 8120, 8122, 8130, 8140, 8211, 8249, and 8255) (1). Enrichment was measured as a binary variable, (1= Yes, statistically higher level of cancer than the general population; 0=No). We annotated each of the 6,416 families as enriched or not enriched for the 17 cancers considered. Familial enrichment was estimated by comparing the observed rate of each cancer type to published rates of cancer incidence in the Utah population assuming a Poisson distribution (16). Families were considered enriched for a certain cancer type if the rate of cancer cases in the family were significantly higher than expected (α≤ 0.05) and flagged accordingly. Sensitivity analyses using a hypergeometric distribution found that the enrichment categorization was equivalent to the categorization using the Poisson distribution.

### Distance Measure and Selection of k

Due to dichotomous values value for enrichment, we used the Hamming distance(17) as our distance metric. This metric is commonly used in natural language processing and information theory for quantifying the similarity/dissimilarity for discrete values. This distance measure has also been used for clustering SNP-sets (18, 19). K was selected by running an iterative set of models from k=2 to k=20 and using silhouette plots to determine the appropriate k (20). In addition to using silhouette plots, we quantified risk of BCa mortality, familial risk of cancer, sex-specific risk of cancer, and enrichment of known BRCA1 and BRCA2 mutation carriers within each cluster using traditional methods.

### Assessment of families after cluster identification

Cox models were used to identify differences in BCa-specific mortality of BCa pivots by FMC cluster. Cause of death was obtained from state-wide death certificate records. Descriptive statistics were generated for overall survival and BCa survival. We excluded 134 pivots for inadequate follow-up information. Age and sex matched controls were used. Duration was measured as time to death or last known date residing in Utah. Individuals who died from non-BCa causes were considered censored at time of death in the BCa-specific survival analyses. We tested for differences in five-year and overall survival between FMCs as well as sex-differences in survival.

To quantify risk in relative types and spouses in each FMC, a case-control design was used. Cancer-free population controls were selected randomly, without replacement, from the UPDB and matched 5:1 to BCa pivots by sex and birth year. Individuals previously excluded due to belonging to a sibset with multiple BCa cases were included in this analysis (NBCa=6,752; NControls=33,752). Relatives of BCa pivots and controls were identified. Relatives with unknown sex were removed (n=35). The case-control analysis included about 2 million relatives (RBCa=345,921; RControls=1,620,791). Spouses (STotal= 32,173; SBCa=5,948; SControls=26,225) were considered a proxy for later life environmental exposures (smoking, etc.). For each FMC, familial risks were estimated using Cox regression for cancer in relatives of BCa pivots compared to the relatives of controls. Hazard risk ratios (HRR) were estimated for FDRs, SDRs, and first cousins together and separately. We tested for sex-specific risks within the FMC clusters via interaction term. All tests were two-sided and statistical significance was considered at p<0.05.

The Utah Cancer Control Program (UCCP) aims to assess the burden of hereditary cancers at the population level by linking the UPDB to BRCA1 and BRCA2 genetic testing data from medical records, the Utah Cancer Registry, and familial cancer registry data for state-wide surveillance (IRB_00079328). This information was linked to individuals and family members in our BCa analytic files and allowed us to test for the enrichment of BRCA1 and BRCA2 families within clusters. Families were labeled BRCA1 or BRCA2 positive if at least one family member tested positive for a mutation and BRCA negative if all family members tested negative for BRCA1/2. We then tested for a higher proportion of BRCA1, BRCA2, or BRCA negative families per cluster relative to the proportion of BRCA1, BRCA2, or BRCA negative families in the population controls using logistic regression. We consider these results to be descriptive due to selection into the testing pool. In lieu of reporting odds ratio estimates, we report p-values (full results available upon request). This study was approved by the Institutional Review Board of the University of Utah (IRB_00088870).

## Results

We identified 6,752 BCa pivots, 345,921 relatives and 5,948 spouses (Table 1). The number of family members ranged from 10 to 334. Median age at diagnosis of BCa pivots ranged from 66.9 to 72.7 years and 5-year BCa-specific mortality ranged from 7.98% to 11.82%. We found no statistically significant between-cluster or sex-specific differences in mortality (Table 2).

**Table 1:** Number of Bladder Cancer (BCa) Pivots and Relatives

| Cluster | Total Number<br>of BCa<br>Pivots/Families | Number<br>of<br>Relatives | Number<br>of<br>Spouses |
| --- | --- | --- | --- |
| 1 | 531 | 40,537 | 463 |
| 2 | 338 | 24,393 | 295 |
| 3 | 1758 | 45,612 | 1,585 |
| 4 | 571 | 43,768 | 530 |
| 5 | 411 | 20,135 | 370 |
| 6 | 477 | 25,837 | 416 |
| 7 | 525 | 39,251 | 555 |
| 8 | 289 | 20,705 | 243 |
| 9 | 326 | 19,292 | 287 |
| 10 | 529 | 21,572 | 445 |
| 11 | 679 | 33,091 | 577 |
| 12 | 208 | 11,728 | 182 |
| Total | 6752 | 345,921 | 5,948 |

\*Only included if 10+ familial BCa cases in cluster

**Table 2.** Characterization of the pivot bladder cancer cases by familial multi-cancer configuration (FMC)

| **FMC** | **Pivot**<br>**Average**<br>**Age at**<br>**Diagnosis** | **N<sup>Pivots</sup>**<br>**Mortality**<br>**Analysis** | **BCa**<br>**Mortality 5**<br>**years** | **BCa**<br>**Mortality** |
| --- | --- | --- | --- | --- |
| 1 | 72.67 | 520 | 10.77% | 13.65% |
| 2 | 72.61 | 326 | 7.98% | 10.74% |
| 3 | 66.89 | 1737 | 8.92% | 10.94% |
| 4 | 72.3 | 553 | 9.22% | 10.85% |
| 5 | 68.1 | 408 | 9.80% | 12.25% |
| 6 | 68.02 | 468 | 10.68% | 13.68% |
| 7 | 70.71 | 525 | 10.72% | 12.96% |
| 8 | 70.47 | 281 | 10.32% | 15.30% |
| 9 | 69.87 | 318 | 8.49% | 11.01% |
| 10 | 68.32 | 520 | 9.81% | 11.54% |
| 11 | 69.63 | 659 | 10.93% | 13.20% |
| 12 | 69.29 | 203 | 11.82% | 14.78% |
| Total | 71.07 | 6518 | 9.79% | 12.18% |

### Familial multi-cancer configurations

We found 12 distinct FMCs (FMC1-12) using k-medioid clustering; each exhibited different patterns of cancer aggregation (Figure 2 and Supplementary Figure 1). Four of the FMCs represented families enriched for BCa and other cancers (FMC4, FMC1, FMC8, and FMC6; nfamilies=1868, 28.1%). These four clusters were the focus of our subsequent analyses. The other eight FMCs were not enriched for BCa and not considered ‘familial BCa clusters’. Seven were enriched only for other cancers, which could represent shared genetic and environment risks. One (FMC3, n=1758; 26.4%) indicated no familial enrichment of cancer and was considered representative of sporadic BCa. Further characterization of the non-familial and sporadic BCa clusters is found in the Supplementary Material.

**Figure 2.**
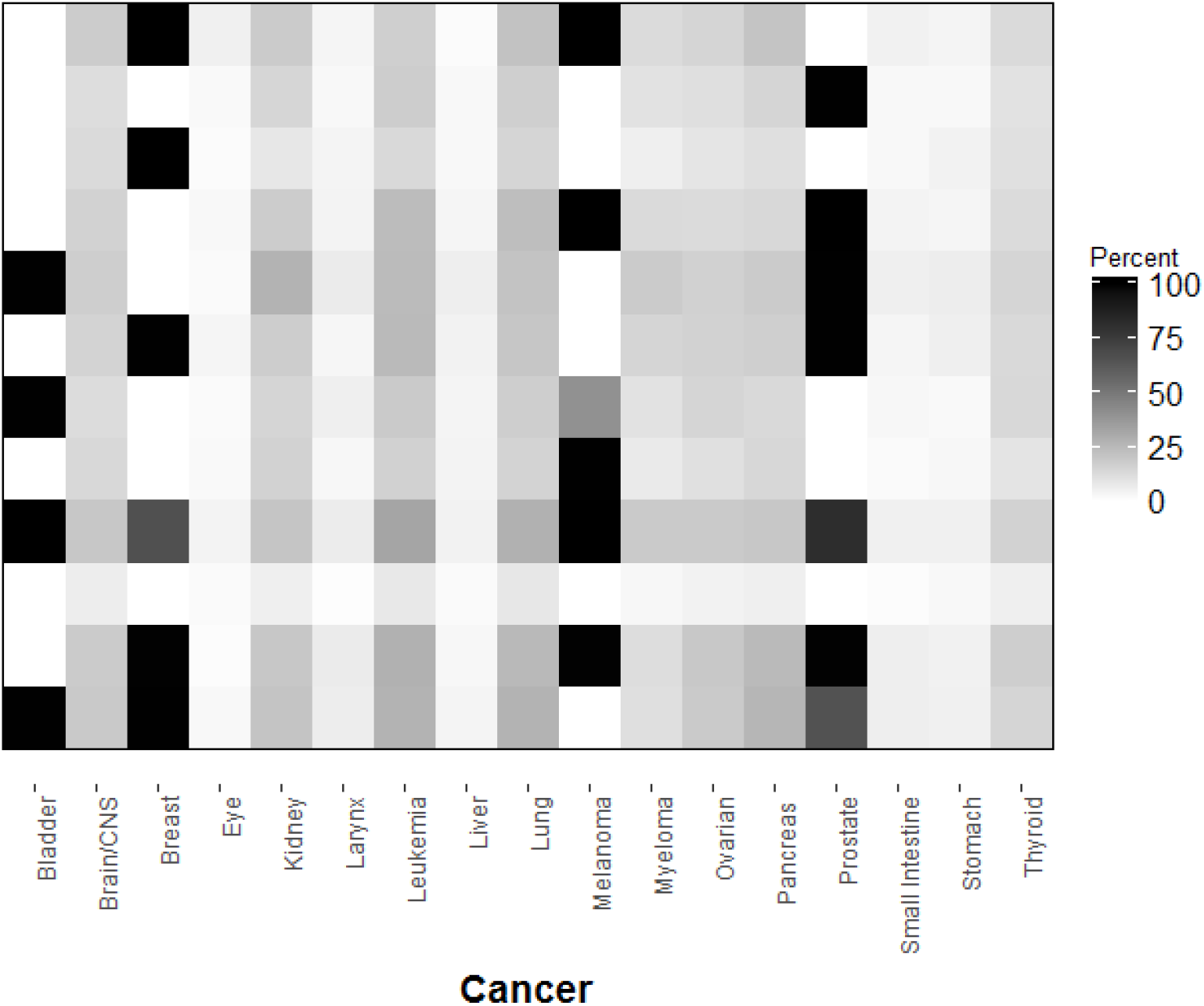
The percent of families enriched for cancer in each Familial Multi-Cancer (FMC) cluster.

The proportion of BCa families captured by the four familial BCa FMCs were 8.5%, 7.9%, 4.3%, and 7.1% for FMC4, FMC1, FMC8, and FMC6, respectively. Figure 3a shows the heat-plot for these FMCs, indicating their different patterns of enrichment. Enriched families are light blue (a statistically higher rate than the population). Of the 17 cancer types studied, breast, melanoma, and prostate were most commonly enriched in the familial BCa clusters. Stomach, liver, larynx, small intestine, and ocular cancers were less commonly enriched.

**Figure 3.**
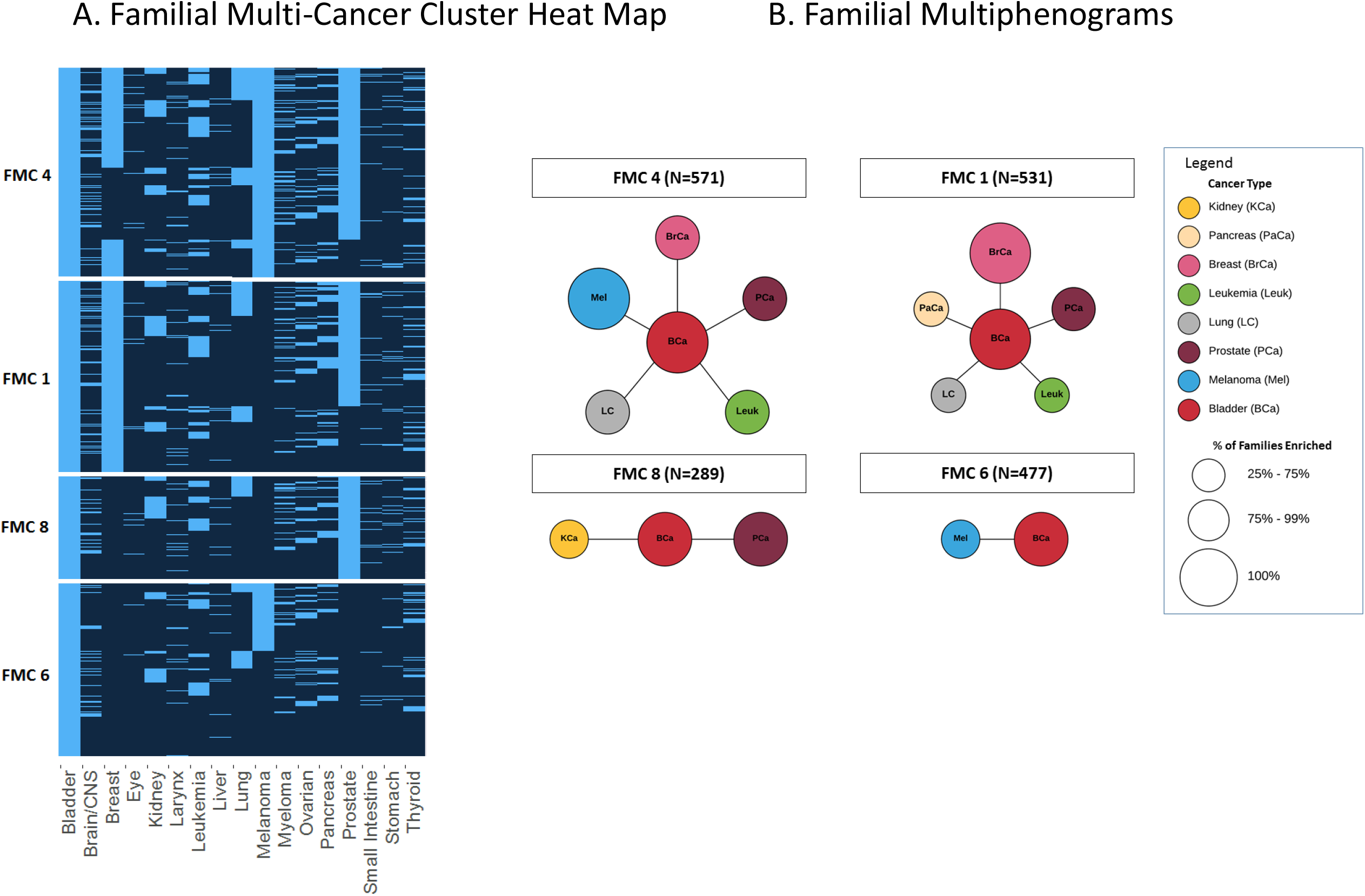
Panel A. Familial multi-cancer configurations for high risk bladder cancer (BCa) families. The y-axis groups each family (row) by FMC and the x-axis shows the type of cancer. Cells are highlighted in light blue if the number of cancers in the family is higher than expected given the population rate. Panel B. Familial Multiphenograms illustrating the four familial BCa multi-cancer configurations

We display a multiphenogram for each FMC in Figure 3b. We defined an FMC as “strongly enriched” if at least 25% of families in the FMC was enriched. FMC4 was strongly enriched for BCa, breast, melanoma, prostate, lung and leukemia (Figure 3). Families in FMC1 were strongly enriched for BCa, pancreas, lung, leukemia, and breast cancer. Families in FMC8 were strongly enriched for BCa, prostate and kidney cancer. Families in FMC6 were strongly enriched for BCa and melanoma.

### Relative-Specific Risks in Familial BCa FMCs

Supplementary Tables 1 - 7 and Supplementary Figure 2 show HRRs by sex and relative type for each cancer within familial BCa FMCs across all families in the FMC compared to control families. This analysis quantified familial risk and provided separate HRRs for relative types and sex. Consistent with our designation of ‘familial BCa’ FMC clusters, relatives in FMC4, FMC1, FMC8, and FMC6 had more than twice the risk of BCa than relatives of controls and FMC6 had the highest risk (HRRFMC6=4.02, p<0.0001).

The FMC4 multiphenogram indicates aggregation with BCa, melanoma, breast, prostate, leukemia and lung cancer (Figure 3B). Relatives of BCa pivots in FMC4 have more than twice the risk of BCa compared to relatives of controls (HRR=2.49, p<0.0001). FMC4 is also characterized by an increased risk of melanoma (HRR=2.14, p<0.0001), breast (HRR=1.12, p=0.0002), prostate (HRR=1.10, p=0.0001). While more than 25% of families are enriched for leukemia and lung cancer, risk for the whole cluster does not reach statistical significance when controlling for birth year and sex (HRRluekemia=1.13, p=0.07; HRRlung= 1.02, p=0.75).

The FMC1 multiphenogram indicates aggregation with BCa, breast, prostate, pancreas, lung cancer and leukemia (Figure 3B). Relatives of BCa pivots in FMC1 had more than twice the risk of BCa than relatives of controls (HRR=2.49, p<0.0001). FDR, SDR, and first cousins were at increased risk for breast cancer (overall HRR=1.44, p<0.0001). FDR were at increased risk for prostate (HRR=1.22, p=0.007). Lung cancer was nominally increased in relatives overall (HRR=1.13, p=0.01). While more than 25% of families were enriched for pancreatic cancer and leukemia, risk for the entire FMC did not reach statistical significance when controlling for birth year and sex (HRRpancreas=1.07, p=0.39; HRRleukemia=1.36, p=0.54). Additionally, FDR and SDR in this FMC were at increased risk for thyroid cancer (HRRFDR=1.74, HRRSDR=1.65).

The FMC8 multiphenogram indicates strong aggregation for BCa, prostate, and kidney cancers (Figure 3B). Relatives of BCa pivots were found to be nearly three-times more likely to be diagnosed with BCa than relatives of controls (HRR=2.71, p<0.0001), and this risk remained reasonably consistent by relative type (HRRFDR=2.74, p<0.0001; HRRSDR=3.43, p<0.0001; HRRFC=2.43, p<0.0001). Relatives had increased risk of prostate cancer (HRR=1.29, p<0.0001) and kidney cancer (HRR=1.40, p=0.001). We also found nominal evidence for increased risk of lung cancer (HRR=1.16, p=0.04) and myeloma (HRR=1.35, p=0.02).

The FMC6 multiphenogram indicates strong aggregation with BCa and melanoma (Figure 3B). Consistent with those findings, we found significant increased risk for BCa and melanoma (HRRBCa=2.71, p<0.0001; HRRmelanoma=1.21, p=0.002).

### Adult Environmental Exposure

Increased risk of cancer in spouses suggests adulthood exposure to an environmental toxin that may explain some patterns of clustering within an FMC. Spouses of BCa cases in FMC4 had increased risk of prostate (HRR=1.65, p=0.03) and kidney (HRR=2.33, p=0.04) cancers. Spouses of BCa pivots in FMC6 were at increased risk of kidney (HRR=2.76, p=0.03) and larynx (HR=11.4, p=0.0014) cancers. There was no difference in risk for spouses of BCa cases in FMC1 or FMC8. Adult environmental exposures may not play a major role in cancer risk for these clusters.

### Sex-Specific BCa Risk

Sex-specific results can be found in Supplementary Figure 1 and Supplementary Tables 2 and 3. Significant difference in sex-specific BCa risk were observed for relatives in FMC4 and FMC6. In FMC4 the female risk of BCa was significantly greater (HRMaleFMC4=2.31 and HRFemaleFMC4=3.15, sex-difference p=0.0002). In FMC6, while risk for BCa is high in both sexes compared to controls, the female-specific risk was significantly lower (HRMaleFMC6=4.22 and HRFemaleFMC6=3.35, sex-difference p=0.035). In addition, the risk of melanoma, lung, and myeloma varied by sex in FMC6, with females having a higher risk for melanoma and lower risk of lung and myeloma compared to males in these families. In FMC1, female relatives were found at increased risk for brain/CNS cancers (HRRFMC1=1.22, p=0.07; sex-difference p=0.005), while male relatives are at decreased risk for brain/CNS cancers (HRRFMC1=0.71, p=0.03).

### Reduced Cancer Risk

While the initial family-based clustering algorithm did not consider decreased cancer risk, when assessing relative-specific risks we identified patterns of reduced risk by cancer type within each FMCs (Supplementary Table 1). There is a lower risk for pancreatic cancer (HRR=0.77, p=0.003) in FMC4, melanoma (HRR=0.20, p<0.0001) in FMC1, breast cancer (HRR=0.29, p<0.0001) and melanoma (HRR=0.19, p<0.0001) in FMC8, and breast (HRR=0.36, p<0.0001) and prostate cancer (HRR=0.27, p<0.0001) in FMC6.

### BRCA Families

BRCA1/2 genetic test results were available for 518 individuals from 257 families. Families were classified as BRCA1 or BRCA2 families if at least one individual tested positive for the mutation and BRCA negative if no individuals in the family tested positive for BRCA1/2. Two families had carriers of BRCA1 and BRCA2 and were placed in both categories (NBRCA1=80, NBRCA2=62, NNeg=117). We found significant enrichment of BRCA1 families in one of the familial BCa clusters (FMC4 p<0.001) and two non-familial clusters (FMC2 p=0.008 and FMC12 p=0.02). FMC1 was enriched with BRCA2 families (p=0.03). Two families were sigificantly enriched for BRCA1/2 negative families, indicaticating that they have familial risk of cancer that qualifies them for testing, but is not related to the BRCA1/2 genes (FMC7 p=0.03 and FMC12 p=0.007).

## Discussion

We have described familial multi-cancer configurations (FMC), illustrating 12 patterns of multi-cancer clustering surrounding BCa patients. Four FMCs exhibited strong familial risk of BCa, as well as co-aggregation with other cancer types. Eight FMCs did not have increased risk of BCa (non-familial BCa). The largest of these did not show increased risk to any cancer and was considered representative of sporadic BCa. We also identified risk variation by sex and relation type across FMCs. Patterns of multi-cancer risk in spouses of BCa cases suggests some FMCs may have environmental risk factors. The four different FMC clusters identified for BCa illustrate the potential of our network-inspired approach to simultaneously assess multiple cancer risks, shifting focus away from a unidimensional definition of family history to a more comprehensive view of family history and risk prevention.

Enrichment of BRCA1 families in FMC4 and BRCA2 families in FMC1 provides support for the utility of a multi-cancer configurations for identifying families genetically predisposed to cancer and the possible pleiotropic effects of genetic mutations. BRCA1/2 genes function across multiple tissue types and development of cancerous cells is not constrained to breast and ovarian tissue. BRCA1 and BRCA2 founder mutations have been associated with prostate and pancreatic cancer risk (21-23). There is an increased risk of breast and prostate cancer in FMC4 and FMC1. FMC1 has an increased risk of pancreatic cancer, which is consistent with reports that pancreatic cancer risk is higher for BRCA2 relative to BRCA1 mutation carriers(24). FMC4 has elevated risk of melanoma, which is consistent with significantly elevated risk of melanoma in BRCA1 families (25).

The concept of phenomes, extending the phenotype to include multiple disease types to characterize genotype-phenotype relationships, is not new. However, the phenome is usually referred to as the set of all phenotypes within an individual. Recent studies demonstrated the feasibility of ‘phenome-wide association scans’ (PheWAS), using genetic data linked to medical records to identify multiple phenotypes associated with a single genotype (26). Pan-cancer analyses are another familiar concept. The Cancer Genome Atlas (TCGA) launched the Pan-cancer analysis project in 2012 with the goal of combining molecular data across large numbers of tumor types to compare -omics data across tumors, potentially allowing for the identification of thematic pathways (27). However, these are independent individuals. Here we utilized family-based data to investigate familial patterns of pan-cancer clustering, a novel familial extension to pan-cancer studies.

Molecular diagnostics and cancer-specific subtypes are becoming an essential component of clinical decision making, however current approaches are typically organ-specific or do not include context for understanding the interplay between genes and environoment. The most recent BCa-specific analysis of TCGA data found five distinct subtypes; 1) luminal-papillary, 2) luminal-infiltrated, 3) luminal, 4) basal-squamous, and 5) neuronal (28). Other studies have shown these subtypes closely align to cancers in other organs. The PAM50 algorithm (originally developed for breast cancer) is used to classify breast, prostate, bladder and lung cancer (29-32), evidence that molecular subtypes may share common etiologies. Subtypes of BCa cancer have clinically meaningful differences (33), however what predisposes an individual to a particular subtype is unknown. Understanding etiology of the disease has clinical potential because it allows for increased screening in at-risk populations and may guide treatment decisions at early stages of diagnosis. Combining familial multi-cancer phenotypes, as we developed here, with tumor molecular data has potential for identification and characterization of BCa subtypes that may share common etiological tumorigenic pathways.

Many genetic and environmental risk factors have been proposed in BCa risk, which could manifest as different multi-cancer configurations across a spectrum of organs. FDRs likely share similar environments throughout the life course and therefore familial aggregation of BCa cancer may not be entirely genetic. For example, family members exposed to arsenic through common drinking wells during childhood may have later life risk for bladder and other related cancers (34). Prostate and kidney risk cancer increases with arsenic exposure (35, 36) and FMC8 had increased risk of both. Genetic predispositions may also make individuals sensitive to environmental exposures. For example, arsenic metabolism may vary between individuals and the rate of metabolism affects risk for adverse health outcomes (37). Moreover, individuals with N-acetyltransferase 2 (NAT2) slow acetylator and glutathione S-transferase µ1 (GSTM1)-null genotypes may have increased risk for BCa when exposed to carcinogens through smoking or occupational risk (38-40).

There are limitations with our method of familial multi-cancer clustering discovery. First, the clustering method did not factor in ages of the pedigree members and therefore some families may have younger members on average than others. However, our subsequent analyses (comparing familial cancer risk to population controls) did control for age and sex, and hence we feel our results of distinct multi-cancer configuriations is robust. Second, this method did not factor in age at diagnosis or histopathological information. Future versions of this method would be strengthened by considering those factors. Third, this method did not utilize all information in pedigree structure and future studies should test methods that take advantage of that information. Despite these limitations, application of this approach could provide important insight to numerous cancers and other age-related chronic diseases.

## Conclusions

This study identified four BCa FMCs with different patterns of BCa risk by age, sex, and relative type. Additionally, we showed unique association patterns with cancers of other organs. The strongest risk was for prostate, melanoma, and breast cancers.

## Data Availability

Data for this study are not publicly available due to privacy restrictions.

## References

1. Martin C, Leiser CL, O’Neil B, Gupta S, Lowrance WT, Kohlmann W, et al. Familial cancer clustering in urothelial cancer: a population-based case-control study. J Natl Cancer Inst 2018; 110(5):527–533.

2. Yu H, Hemminki O, Försti A, Sundquist K, Hemminki K. Familial urinary bladder cancer with other cancers. Eur Urol Oncol 2018; 1(6): 461–466.

3. Bermejo JL, Sundquist J, Hemminki K. Sex-specific familial risks of urinary bladder cancer and associated neoplasms in Sweden. Int J Cancer 2009;124(9):2166–2171.

4. Hemminki K, Sundquist J, Brandt A. Do discordant cancers share familial susceptibility? Eur J Cancer 2012;48(8):1200–1207.

5. Frank C, Fallah M, Sundquist J, Hemminki A, Hemminki K. Population landscape of familial cancer. Sci Rep 2015;5:12891.

6. Teerlink CC, Albright FS, Lins L, Cannon-Albright LA. A comprehensive survey of cancer risks in extended families. Genet Med 2012;14(1):107–114.

7. Mérette C, King MC, Ott J. Heterogeneity analysis of breast cancer families by using age at onset as a covariate. Am J of Hum Genet 1992;50(3):515–519.

8. Hall JM, Lee MK, Newman B, Morrow JE, Anderson LA, Huey B, et al. Linkage of early-onset familial breast cancer to chromosome 17q21. Science 1990;250(4988):1684–1689.

9. Easton DF, Bishop DT, Ford D, Crockford GP. Genetic linkage analysis in familial breast and ovarian cancer: Results from 214 families. The Breast Cancer Linkage Consortium. Am J Hum Genet. 1993;52(4):678–701.

10. Samadder N, Smith K, Hanson HA, Pimentel R, Wong J, Boucher K, et al. Familial risk in patients with carcinoma of unknown primary. JAMA Oncol. 2016;2(3):340–346.

11. Anderson RE, Hanson HA, Patel DP, Johnstone E, Aston KI, Carrell DT, et al. Cancer risk in first- and second-degree relatives of men with poor semen quality. Fertil Steril 2016; 106(3):731–738.

12. DuVall SL, Fraser AM, Rowe K, Thomas A, Mineau GP. Evaluation of record linkage between a large healthcare provider and the Utah Population Database. J Am Med Inform Assoc 2012;19(e1):e54–59.

13. Edelman LS, Guo J-W, Fraser A, Beck SL. Linking clinical research data to population databases. Nurs Res 2013;62(6):438–444.

14. DerKinderen DJ, Koten JW, Nagelkerke NJ, Tan KE, Beemer FA, Den Otter W. Non-ocular cancer in patients with hereditary retinoblastoma and their relatives. Int J Cancer 1988;41(4):499–504.

15. Park H-S, Jun C-H. A simple and fast algorithm for K-medoids clustering. Expert Syst Appl 2009;36(2, Part 2):3336–3341.

16. Sweeney C, Herget KA, Otto VY, McFadden S, Millar MM. Cancer in Utah: an overview of incidence and mortality 2004 - 2013. Utah Cancer Registry; 2016.

17. Hamming RW. Error detecting and error correcting codes. Bell System Technical Journal. 1950;29(2):147–160.

18. Wang C, Kao W-H, Hsiao CK. Using Hamming distance as information for SNP-Sets clustering and testing in disease association studies. PLoS One 2015;10(8):e0135918.

19. Imai A, Nakaya A, Fahiminiya S, Tetreault M, Majewski J, Sakata Y, et al. Beyond homozygosity mapping: family-control analysis based on Hamming distance for prioritizing variants in exome sequencing. Sci Rep 2015;5:12028.

20. Rousseeuw PJ. Silhouettes: A graphical aid to the interpretation and validation of cluster analysis. J Comput Appl Math 1987;20:53–65.

21. Agalliu I, Gern R, Leanza S, Burk RD. Associations of high-grade prostate cancer with BRCA1 and BRCA2 founder mutations. Clin Cancer Res 2009; 15(3):1112–1120.

22. Cavanagh H, Rogers KM. The role of BRCA1 and BRCA2 mutations in prostate, pancreatic and stomach cancers. Hered Cancer Clin Pract 2015;13(1):16.

23. Stadler ZK, Salo-Mullen E, Patil SM, Pietanza MC, Vijai J, Saloustros E, et al. Prevalence of BRCA1 and BRCA2 mutations in Ashkenazi Jewish families with breast and pancreatic cancer. Cancer 2012;118(2):493–499.

24. Risch HA, McLaughlin JR, Cole DE, Rosen B, Bradley L, Fan I, et al. Population BRCA1 and BRCA2 mutation frequencies and cancer penetrances: a kin–cohort study in Ontario, Canada. J Natl Cancer Inst 2006;98(23):1694–1706.

25. Gumaste PV, Penn LA, Cymerman RM, Kirchhoff T, Polsky D, McLellan B. Skin cancer risk in BRCA1/2 mutation carriers. Br J Dermatol 2015;172(6):1498–1506.

26. Denny JC, Ritchie MD, Basford MA, Pulley JM, Bastarache L, Brown-Gentry K, et al. PheWAS: demonstrating the feasibility of a phenome-wide scan to discover gene–disease associations. Bioinformatics 2010;26(9):1205–1210.

27. The Cancer Genome Atlas Research Network, Chang K, Creighton CJ, Davis C, Donehower L, Drummond J, et al. The Cancer Genome Atlas Pan-Cancer analysis project. Nat Genet 2013;45(10): 1113–1220.

28. Robertson AG, Kim J, Al-Ahmadie H, Bellmunt J, Guo G, Cherniack AD, et al. Comprehensive molecular characterization of muscle-invasive bladder cancer. Cell 2018;171(4):1033.

29. Zhao SG, Chang SL, Erho N, Yu M, Lehrer J, Alshalalfa M, et al. Associations of luminal and basal subtyping of prostate cancer with prognosis and response to androgen deprivation therapy. JAMA Oncol 2017;3(12):1663–1672.

30. Damrauer JS, Hoadley KA, Chism DD, Fan C, Tiganelli CJ, Wobker SE, et al. Intrinsic subtypes of high-grade bladder cancer reflect the hallmarks of breast cancer biology. Proc Natl Acad Sci U S A 2014;111(8):3110–3115.

31. Parker JS, Mullins M, Cheang MCU, Leung S, Voduc D, Vickery T, et al. Supervised risk predictor of breast cancer based on intrinsic subtypes. J Clin Oncol 2009;27(8):1160–1167.

32. Siegfried JM, Lin Y, Diergaarde B, Lin H-M, Dacic S, Pennathur A, et al. Expression of PAM50 genes in lung cancer: evidence that interactions between hormone receptors and HER2/HER3 contribute to poor outcome. Neoplasia 2015;17(11):817–825.

33. Seiler R, Winters B, Douglas J, van Rhijn BWG, Sjödahl G, Lerner SP, et al. Muscle-invasive bladder cancer: molecular subtypes and response to neoadjuvant chemotherapy. J Clin Oncol 2017;35(6_suppl):281–281.

34. Fernández MI, López JF, Vivaldi B, Coz F. Long-term impact of arsenic in drinking water on bladder cancer health care and mortality rates 20 years after end of exposure. J Urol 2012;187(3):856–861.

35. Benbrahim-Tallaa L, Waalkes MP. Inorganic arsenic and human prostate cancer. Environ Health Perspect 2008;116(2):158–164.

36. Ferreccio C, Smith AH, Durán V, Barlaro T, Benítez H, Valdés R, et al. Case-control study of arsenic in drinking water and kidney cancer in uniquely exposed Northern Chile. Am J Epidemiol 2013;178(5):813–8.

37. Smith AH, Steinmaus CM. Health effects of arsenic and chromium in drinking water: recent human findings. Annu Rev Public Health 2009;30(1):107–122.

38. Moore LE, Baris DR, Figueroa JD, Garcia-Closas M, Karagas MR, Schwenn MR, et al. GSTM1 null and NAT2 slow acetylation genotypes, smoking intensity and bladder cancer risk: results from the New England bladder cancer study and NAT2 meta-analysis. Carcinogenesis 2011;32(2):182–189.

39. Krech E, Selinski S, Blaszkewicz M, Burger H, Kadhum T, Hengstler JG, et al. Urinary bladder cancer risk factors in an area of former coal, iron, and steel industries in Germany. J Toxicol Environ Health A 2017; 80(7-8):430-438.

40. Ma C, Gu L, Yang M, Zhang Z, Zeng S, Song R, et al. rs1495741 as a tag single nucleotide polymorphism of N-acetyltransferase 2 acetylator phenotype associates bladder cancer risk and interacts with smoking: a systematic review and meta-analysis. Medicine (Baltimore) 2016;95(31):e4417.

